# Questioning the Seasonality of SARS-COV-2: A Fourier Spectral Analysis

**DOI:** 10.1101/2022.01.26.22269886

**Authors:** Riccardo Cappi, Luca Casini, Davide Tosi, Marco Roccetti

**Author notes:** **Corresponding Author:** Luca Casini.

## Abstract

**Objectives:** COVID-19 recurrent waves have ignited a debate on the role of seasonality in the contagion resurgence. Two opposite positions emerged. Those convinced of a sinusoidal seasonality recurring over one-year period driven by climate. Those believing that a fluctuation of the outbreaks repeats without limits, in the absence of control measures. We studied the series of daily confirmed SARS-CoV-2 cases in 30 different countries (February 2020 - December 2021), investigating the hypothesis of whether seasonal and geographic variations may guide the pandemic trajectory.

**Methods:** We chose 30 countries from different geographies and climates. With a discrete Fourier transform, we performed a spectral analysis of the series of the daily SARS-CoV-2 infections, looking for peaks in the frequency spectrum that could correspond to a recurrent cycle of a given length. This analysis allows to question whether COVID-19 outbreaks have an observable seasonal periodicity.

**Results:** All the 30 investigated countries see the recurrence of at least one COVID-19 wave, repeating over a period in the range 3 - 9 months, with a peak of magnitude at least half as large as that of the highest peak ever experienced since the beginning of the pandemic until December 2021. We also computed the distance in days between the two higher peaks in each country, and then we averaged those values over the 30 countries, yielding a mean of 190 days (SD 100). This suggests that outbreaks may repeat with cycles of different lengths, without a precisely predictable seasonality of one year.

**Conclusion:** Our findings suggest that COVID-19 outbreaks are likely to occur worldwide, with cycles of repetition of variable lengths. Our Fourier analysis with 30 different countries has not found evidence in favor of a seasonality that recurs over one-year period, solely or with a precisely fixed periodicity.

**Design:** Not Applicable

**Setting:** Not Applicable

**Participants:** Not Applicable

**Interventions:** Not Applicable

**Primary and Secondary Outcome Measures:** Not Applicable

**Article Summary:** *Strengths and Limitations of this Study:*

- The use of a discrete Fourier transform offers the advantage of a temporal decomposition of the time series of the daily SARS-COV-2 cases, allowing to explore the temporal relationships among recurrent outbreaks.
- Analyzing for each country the various repetition cycles of the outbreaks, along with their magnitudes, represents an appropriate method to question the assumption of a seasonal “rise and fall” structure of COVID-19 assumed to recur, almost exclusively, based on a one-year long period.
- We have made inferences about the seasonality of COVID-19 from a purely observational study. The problem has been avoided to quantify, precisely, the role attributable to various climatic factors or control measures, like temperatures or vaccination.
- To take advantage of a natural experiment, the time series of the number of daily SARS-CoV-2 cases were subjected to a discrete Fourier transform without resorting to any form of correction/normalization. The SARS-CoV-2 outbreak (started in December 2021) has not been included in this study as the progression of this wave is still ongoing.
- This analysis presents a natural limitation in time as the pandemic has been in progress so far, for only two years. The Fourier uncertainty principle may render the results at low frequencies somewhat uncertain given this extremely short time domain.

## Introduction

After almost two years from the start of the COVID-19 pandemic, the scientific community is still arguing about many of the characteristics of this virus and its spread, as well as what the best course of action in the fight against it is [1, 2]. While the public may find this lack of consensus disheartening, every scientist knows this was to be expected when dealing with such unprecedented phenomena, especially given the enormous uncertainty around the data that concern it. Every analysis that has tried to leverage information on a global scale, in fact, had to deal with the limitation of having a largely inhomogeneous dataset, where differences in the data collection process and, even more importantly, in the actions taken by each country contributed to confounding the factors being studied [3].

In this complex context, at the end of 2021, as the virus has been around for almost two years, a discussion started about the possibility of COVID-19 following a seasonal pattern, similar to many other viral infections, like measles and flu, for example. This idea gained momentum probably because of how the contagion receded during the summer months in many Western countries, to start climbing back up with the start of autumn and finally reaching a new peak during the winter holidays.

From a scientific perspective, the debate on an infection pattern that repeats over a one-year period has been driven by several analyses investigating the correlation between SARS-COV-2 and various climatic (and environmental) factors, such as temperature, humidity, and UV radiations. The rationale behind this research is that if a negative correlation between SARS-COV-2 and higher temperatures and exposure levels to UV radiation can be demonstrated, then lower COVID-19 infection rates should happen in some given seasons of the year. Humidity, instead, appears to have a U-shaped relation with SARS-CoV-2 infection rates, as only values that hover 50% seem to shorten the virus life.

Along this line of research, remarkable are the studies of D’Amico *et al*., who used a multivariate regression to assess the influence of temperatures and vaccinations on mortality rates in temperate climate countries. They found a negative correlation with temperatures and discovered that the vaccination’s effect grew larger as the temperature decreased [4]. Similarly, Ma *et al*. studied the problem in the United States using a generalized additive mixed model. Their findings are that temperatures are negatively correlated with COVID-19, in an almost linear way, in the range of 20-40° C [5].

However, some other research begins to point out the weaknesses of many of the aforementioned studies. For example, Fontal *et al*., while studying the negative correlation between the virus and both higher temperature and humidity, found that there are moments in which this correlation can be inverted, typically corresponding to summer outbreaks in certain regions [6]. The authors suggest that this can be due when various human activities take over, like intensive use of air conditioning, lack of preventive measures and uncontrolled mass gatherings. Also, Sera *et al*. have expressed their concerns, concluding that the effect of weather, while present, is negligible when compared to the decisive impact of control interventions [7]. More interestingly, Baker et al. have argued that climate factors can play an important factor in the infection when the virus is in the endemic stage. In contrast, during the pandemic stage, it only drives modest changes [8].

Beyond these scientific studies, the positive effect of good weather seems to contrast with several COVID-19 contagions that have broken out, with broad impact, even if with unfavorable climates to the spread of the virus. For example, while this is completely anecdotal, one could wonder what mechanisms were behind the resurgence of the contagion in May 2020 in Israel [9]. Similarly, the 2021 Olympic Games in Japan took place during the summer when the weather was optimal, but the virus spread, even in the presence of high security standards [10]. In the same period, the European Football Championship took place, and this tournament was connected with an increase of new cases in many involved countries [11]. Not to say about the July 4, 2021, US presidential Speech, when the US President declared the final success in beating the pandemic, but a new peak hit strong just a few weeks later [12]. Finally, the third wave across Europe started at the end of 2021 summer in many eastern countries, when the temperatures were still relatively high.

Following this scientific debate, in this work we chose another technical perspective in order to investigate the one-year seasonality hypothesis, employing techniques from signal processing in order to study the presence of evidence towards periodicity in the COVID-19 recurrent outbreaks, or lack thereof. Relying on a Fourier spectral analysis of the daily SARS-CoV-2 infections data, at a worldwide level, we looked for peaks in the frequency spectrum that could inform us about the length of the recurrent cycles of COVID-19 outbreaks. This has allowed us to question on the sinusoidal seasonality assumed to recur over one-year period, solely.

## Methods

We focused our study on a wide selection of worldwide countries, chosen following the Köppen climate classification [13]. This classification divides climates into five main groups, where each group is considered based on seasonal precipitation and temperatures. The five main groups are: tropical (A), dry (B), temperate (C), continental (D) and polar (E). We analyzed 30 different countries that cover all the five groups, with several of the selected countries belonging to two or more groups, given their vast geography (e.g., India, Russia, and the USA, to cite a few). The complete list of the 30 countries follow below, each with its prevalent type of climates: Argentina (B, C), Australia (A, B, C), Austria (D, E), Belgium (C), Brazil (A, C), Canada (C, D, E) Chile (B, C, D), Colombia (A, C), Croatia (C), Denmark (D), France (C), Germany (C, D), Hungary (D), India (A, B, C, D), Indonesia (A), Italy (B, C), Japan (A, C, D), Mexico (A, B, C), Morocco (B, C), Norway (D, E), Portugal (C), Russia (D, E), Saudi Arabia (B), South Africa (B, C), South Korea (C, D), Spain (B, C), Sweden (D, E), Turkey (B, C, D), UK (C) and USA (B, C, D, E).

Notice that our selection includes 18 out of the 20 countries of the Group of 20. China was excluded just because its SARS-COV-2 data are not made available on a regular basis. Also, the European Union (EU) was not considered as a whole. Yet, in place of EU, we included the following EU members: Austria, Belgium, Croatia, Denmark, France, Germany, Hungary, Italy, Portugal, Spain, and Sweden. The period of observation for our study started on February1, 2020 until December 24, 2021, with the decision not to take into consideration the strong SARS-CoV-2 outbreak that hit Europe in December 2021, as the progression of this wave was still ongoing in many of the investigated countries during our analysis.

The method we adopted for our investigation was a Fourier spectral analysis. In particular, we used a Discrete Fourier Transform (DFT) to examine the periods length in the spectrum of the COVID-19 data, by converting the time series of the number of the new daily SARS-CoV-2 cases to the frequency domain [14]. This Fourier frequency spectrum analysis was performed with the precise intent to obtain a converted peak spectrum, indicating the strength and the recurrence of the pandemic waves. In particular, we looked for peaks in the frequency spectrum that could reasonably indicate a periodicity with a certain length. Employing a spectral analysis on the time series of the COVID-19 cases has allowed us to understand, with less ambiguity, the period length of the recurrent outbreaks, instead of counting and observing the number of infections, directly.

The 1-dimensional DFT *y*[*k*] of length *N*, of the length-*N* sequence *x*[*n*], is defined as:

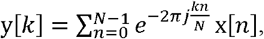

Where *y*[*k*] corresponds to magnitude of the *k*-th frequency and *n* represents the *n*-th day of the time series, with *x* being the daily number of SARS-COV-2 cases registered on that *n*-th day of the series.

It is to notice that the number of the analyzed days (i.e., the sampling period) was equal to 730 (two years). Since our study’s real period of observation started on February 1, 2020 (until December 24, 2021), the string *x* was left padded with zeros to reach 730 samples. This zero padding did not alter the validity of the operation since in all the considered countries no SARS-COV-2 infection was registered before February1, 2020. To conclude, using a Python library called *SciPy (https://scipy.org/)*, we performed a DFT of the time series of the SARS-COV-2 data of each country, that returned all the peaks in the frequency spectrum at their corresponding frequency which can be inverted to obtain the repetition period.

### Patient and Public Involvement

Patients and/or the public were not involved in the design, or conduct, or reporting, or dissemination plans of this research. All the data used are available from the following official site: https://github.com/owid/covid-19-data. Instructions on how to use those data are available at: https://github.com/owid/covid-19-data/blob/master/public/data/README.md. The results of all the DFT computations are fully reproducible by using the method stated above. The authors are open to discussing the code with whoever reaches out by email. The DFT code itself can be downloaded from here: https://github.com/mister-magpie/covid_periodicity.

## Results

Figures 1-5 show the FTs obtained for all the countries subject of our study, using two separate plots. For each country, the leftmost plot reports the time series of the new daily SARS-COV-2 infections during the observed period (February 1, 2020 - December 24, 2021); the rightmost plot shows the result of the Fourier transform applied to the time series of the leftmost plot.

**Figure 1:**
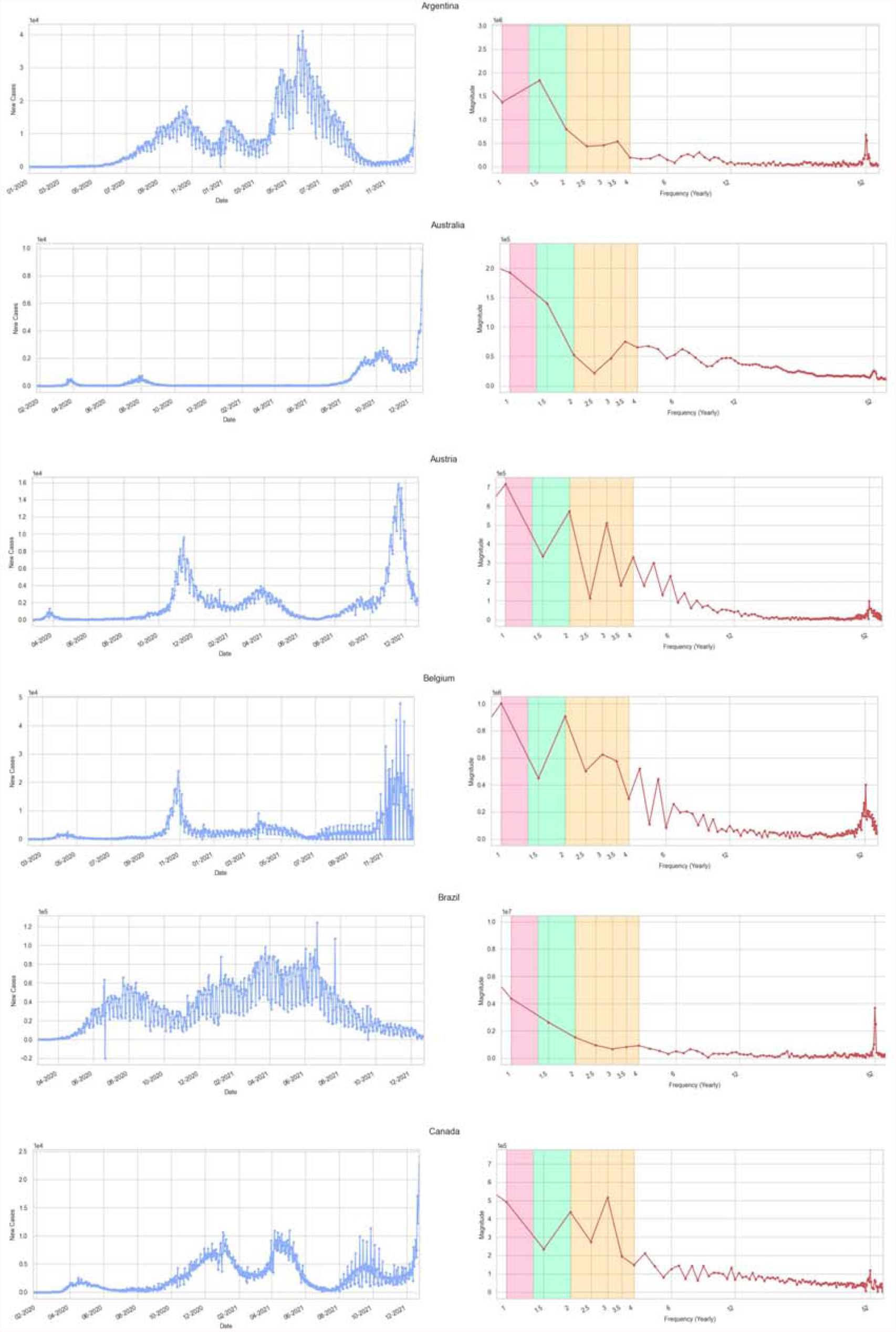
Plots for Argentina, Australia, Austria, Belgium, Brazil, and Canada. Leftmost plots: on the *y* axis number of new daily SARS-CoV-2 cases in the period February1, 2020 - December 24, 2021 (*x* axis). Rightmost plots: Fourier frequency spectrum on the *y* axis, with dots lying on the red line corresponding to wave peaks. Colored sectors indicate precise recurring cycles: pink (9-12 months), green (6-9 months), orange (3-6 months). The *k* index on the *x* axis of the rightmost plots indicated the repetition frequency on a semi-logarithmic scale (on a one-year period).

**Figure 2:**
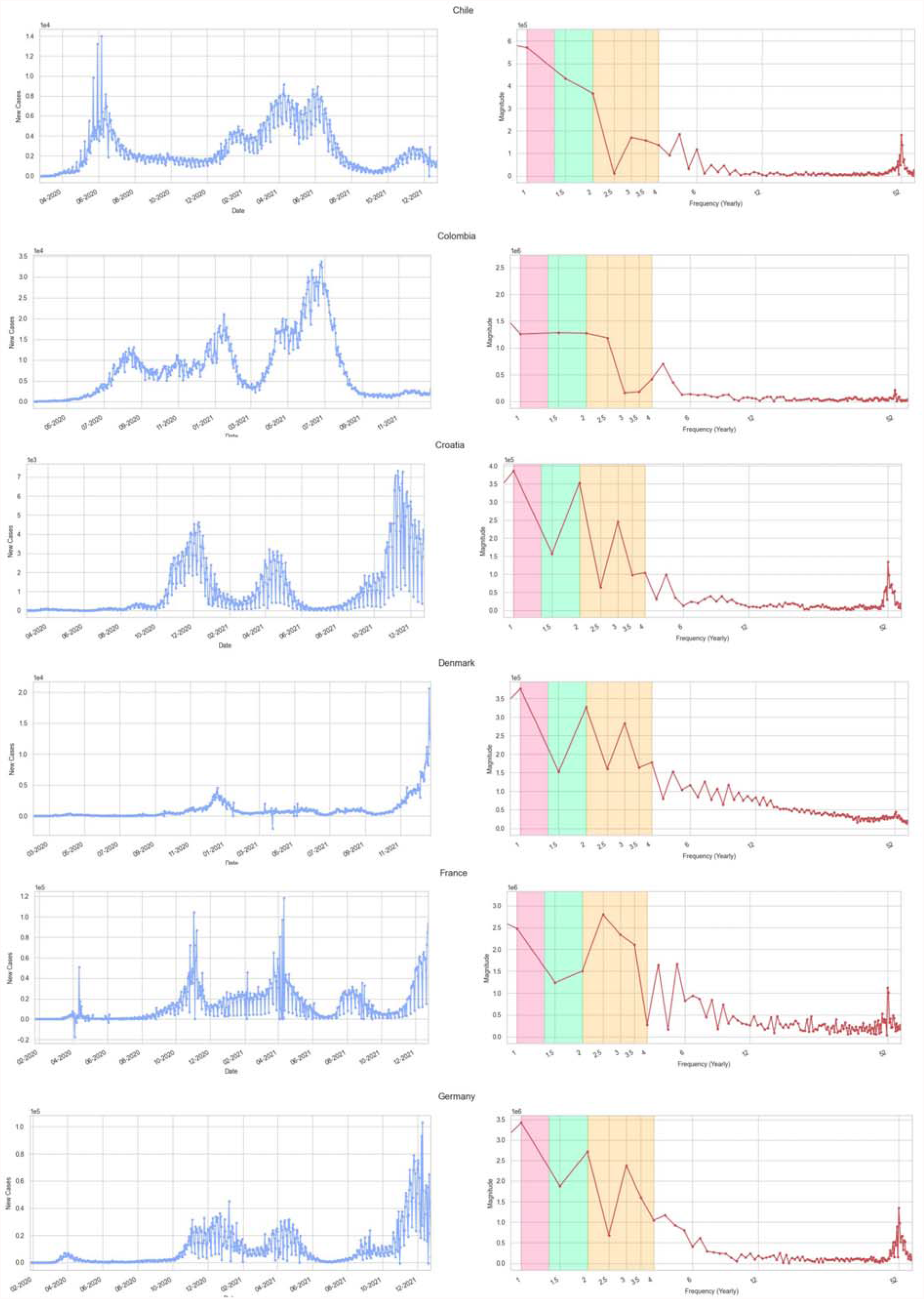
The same plots of Figure 1 but for Chile, Colombia, Croatia, Denmark, France, and Germany.

**Figure 3:**
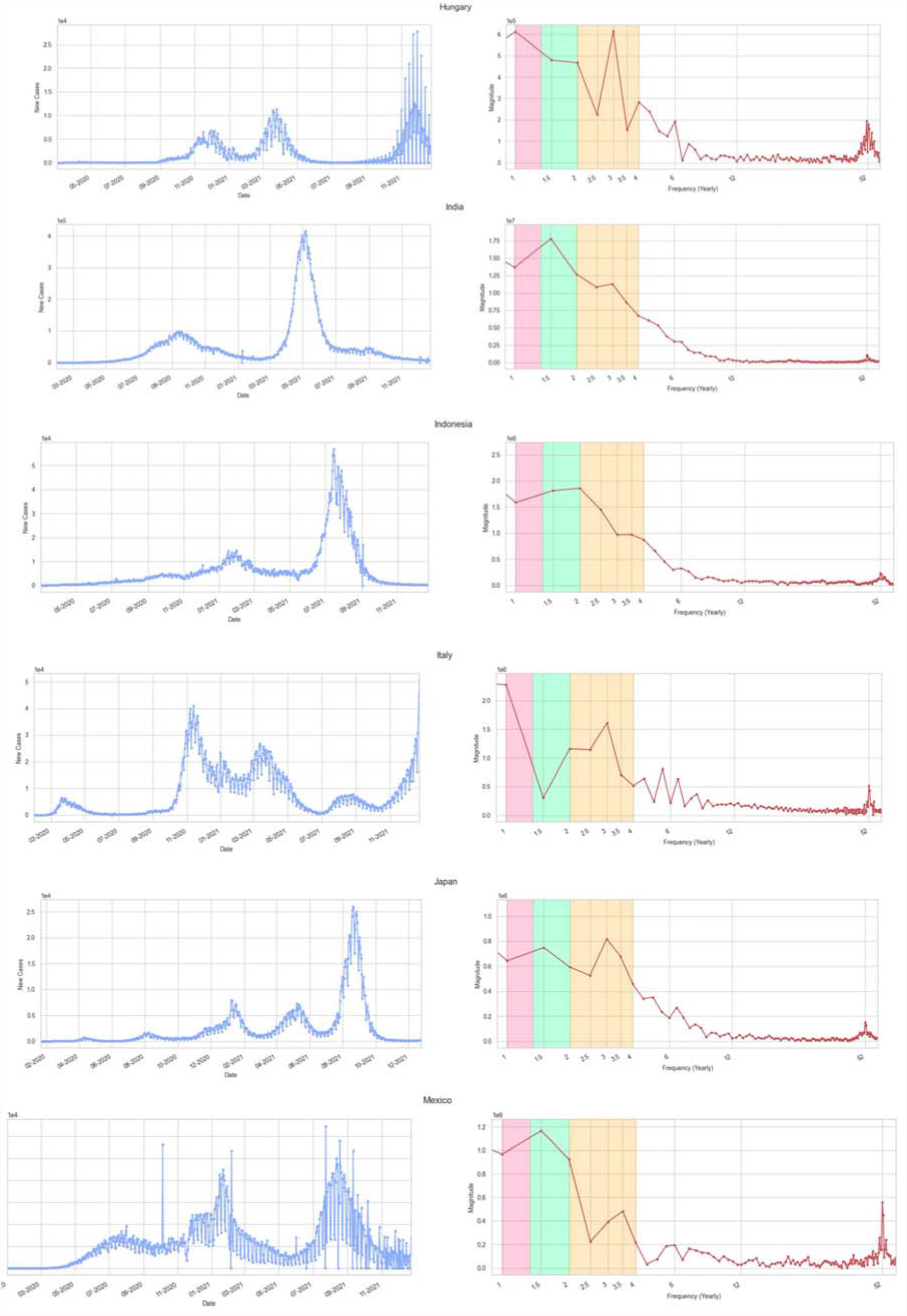
The same plots of Figure 1 but for Hungary, India, Indonesia, Italy, Japan, and Mexico.

**Figure 4:**
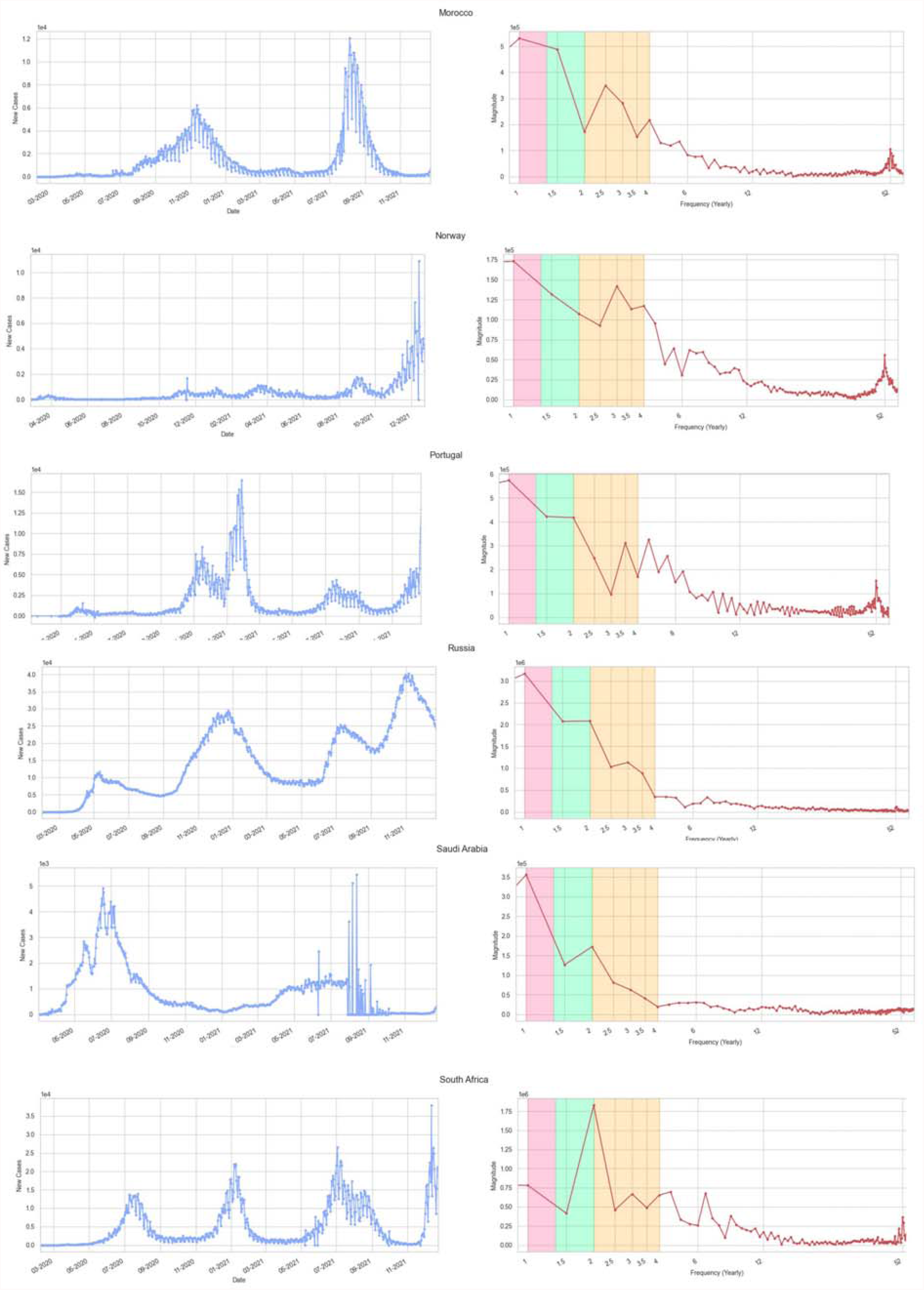
The same plots of Figure 1 but for Morocco, Norway, Portugal, Russia, Saudi Arabia, South Africa.

**Figure 5:**
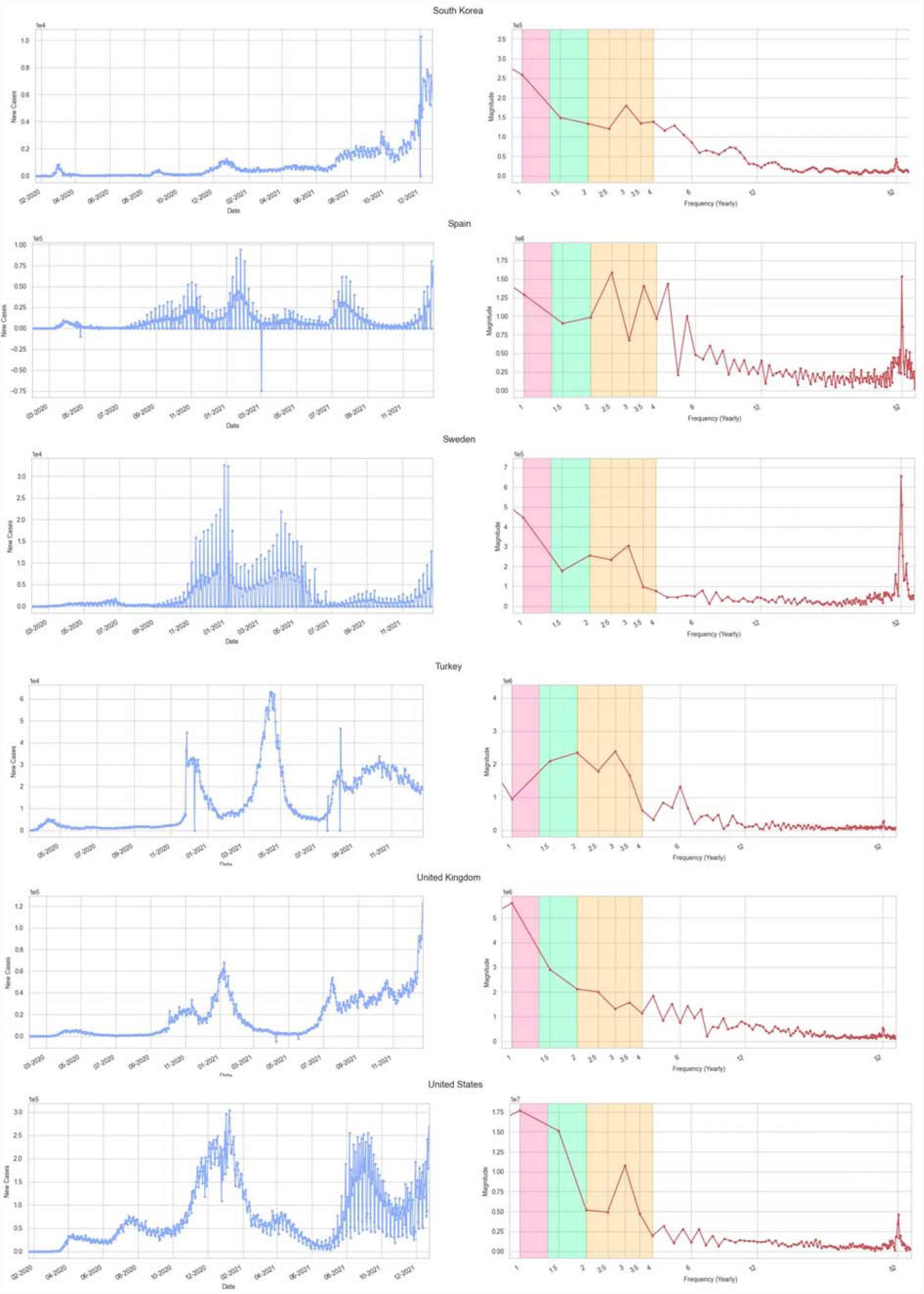
The same plots of Figure 1 but for South Korea, Spain, Sweden, Turkey, UK, and USA.

In all the leftmost plots, *x*[*n*] is the COVID-19 data series of interest, where *x* is the number of daily new infections per each day *n*. All the rightmost plots of Figures 1-5 depict, in the *y* axis, the Fourier frequency spectrum. This spectrum comprises a red line with endpoints at the junctions, representing the COVID-19 peaks. Each peak is depicted with its magnitude (*y* axis) and with its corresponding frequency *k* on the *x* axis, based, in turn, on a semi-logarithmic yearly scale. Two preliminary facts are noteworthy. First, we can observe a peak in the frequency spectrum representing the 7-day cycle associated with the case reporting process, on the rightmost side of all these DFT plots. This was a quite expected fact, since the reporting process causes an oscillation during the week, in almost all the considered countries. Obviously, this peak recurs 52 times in the DFT plot (*k* = 52), being 52 the number of the weeks in one year. Second, we observe a peak with typically the highest magnitude at the opposite side of the spectrum of all our DFT plots (leftmost side). The meaning of this peak is simply that the COVID-19 phenomenon has occurred each year, during the two years of observation, in all the countries of interest. Its repetition cycle is, naturally, equal to 1 (*k* =1).

More interestingly, on the leftmost side of the DFT plots of Figures 1-5 we have three different sectors colored in orange, green and pink (from right to left). Those sectors display temporal intervals, respectively, equal to 3-6 months (orange), 6-9 months (green), and 9-12 months (pink). They should be interpreted as follows: If one observes for a certain country the presence of a peak in a given colored sector of the plot (say the green one, for example), this means that country has been hit by a COVID-19 outbreak, which has recurred with a period of 6-9 months. More precisely, if that peak lies on the *x* axis in correspondence of a value of *k* = 2, this implies that we have had two outbreaks of similar magnitude per year in that country. Coming now to our results, our 30 DFT plots of Figures 1-5 reveal that, in the observed period, all the 30 investigated countries have seen the recurrence of at least one COVID-19 wave, repeating over a variable period in the range 3 - 9 months, with a peak of magnitude (roughly equivalent to the number of new infections) at least half as high as that of the highest peak ever experienced since the beginning of the pandemic until December 2021. These findings suggest that strong COVID-19 outbreaks may repeat with cycles of different lengths, without a precisely predictable seasonality of one year.

Given the well-known Fourier uncertainty principle [15], we developed a further analysis. We returned to the leftmost plots of Figures 1-5, looking for the COVID-19 peaks, recurring in each country, but adopting a more traditional technique. Specifically, using a 7-day rolling average as the raw data of leftmost plots of Figures 1-5 present a weekly periodicity, due to the way COVID-19 tests are carried out and registered, we considered a peak has happened in a given day *n*, if the number of SARS-COV-2 infections registered in that day was larger than the number of daily SARS-COV-2 cases reported in the 28 days both before and after *n*. Not only, but to be considered a peak, the number of infections registered on that day *n* had to be larger than a given threshold computed as the 85% of the average of the daily cases reported in all the days since the beginning of the pandemic until *n*. Choosing 28 days comes from the working definition of wave as provided in [15], where the three quarters of the upward periods of many studied COVID-19 waves lasted less than a month. Similarly, for the downward periods. The rationale behind the concept of having a threshold came, instead, from the need to filter out all the micro peaks. Finally, upon computation of all the peaks for each country during the period of interest, we chose the two highest ones. Then we computed for each such pair the distance in days between them (the Python code for this simple algorithm can be downloaded from: https://github.com/riccardocappi/Seasonality-SARS-CoV-2). Table 1 reports the corresponding results.

**Table 1:** Country, type of climates for that country, number of COVID-19 outbreak peaks, distance in days between the two highest peaks, dates of the two highest peaks, dates of the remaining peaks. The mean distance between the two highest peaks is 190 days (SD 100).

| Country | Climate types | Number of peaks | Distance between the two highest peaks (days) | Dates of the two highest peaks | Dates of the remaining peaks |
| --- | --- | --- | --- | --- | --- |
| Argentina | B/C | 3 | 214 | 2020/10/21<br>2021/05/23 | 2021/01/11 |
| Australia | A/B/C | 3 | 432 | 2020/08/04<br>2021/10/10 | 2020/03/30 |
| Austria | D/E | 5 | 376 | 2020/11/13<br>2021/11/24 | 2020/03/28; 2021/04/01; 2021/09/15 |
| Belgium | C | 4 | 148 | 2020/10/30<br>2021/03/27 | 2020/04/15; 2020/08/12 |
| Brazil | A/C | 4 | 87 | 2021/03/27<br>2021/06/22 | 2020/07/28; 2021/01/12 |
| Canada | C/D/E | 4 | 93 | 2021/01/09<br>2021/04/12 | 2020/04/22; 2021/09/13 |
| Chile | B/C/D | 6 | 55 | 2021/04/14<br>2021/06/08 | 2020/06/12; 2020/10/01; 2021/01/25;<br>2021/11/15 |
| Colombia | A/C | 4 | 159 | 2021/01/20<br>2021/06/28 | 2020/08/16; 2020/11/02 |
| Croatia | C | 6 | 333 | 2020/12/13<br>2021/11/11 | 2020/04/01; 2020/07/15; 2020/08/29;<br>2021/04/21 |
| Denmark | D | 6 | 145 | 2020/12/18<br>2021/05/12 | 2020/04/08; 2020/09/23; 2021/03/16;<br>2021/08/16 |
| France | C | 5 | 161 | 2020/11/07<br>2021/04/17 | 2020/04/18; 2021/02/11; 2021/08/16 |
| Germany | C/D | 4 | 123 | 2020/12/23<br>2021/04/25 | 2020/04/02; 2021/09/10 |
| Hungary | D | 3 | 113 | 2020/12/03<br>2021/03/26 | 2020/04/13 |
| India | A/B/C/D | 2 | 234 | 2020/09/16<br>2021/05/08 | / |
| Indonesia | A | 3 | 167 | 2021/02/01<br>2021/07/18 | 2020/09/26 |
| Italy | B/C | 5 | 126 | 2020/11/16<br>2021/03/22 | 2020/03/26; 2021/01/11; 2021/08/27 |
| Japan | A/C/D | 5 | 226 | 2021/01/11<br>2021/08/25 | 2020/04/15; 2020/08/09; 2021/05/14 |
| Mexico | A/B/C | 4 | 213 | 2021/01/21<br>2021/08/22 | 2020/08/01; 2020/10/09 |
| Morocco | B/C | 4 | 266 | 2020/11/17<br>2021/08/10 | 2020/04/22; 2020/06/25 |
| Norway | D/E | 6 | 167 | 2021/03/22<br>2021/09/05 | 2020/03/29; 2020/11/23; 2021/01/10;<br>2021/05/26 |
| Portugal | C | 5 | 70 | 2020/11/19<br>2021/01/28 | 2020/04/03; 2020/07/13; 2021/07/23 |
| Russia | D/E | 4 | 315 | 2020/12/26<br>2021/11/06 | 2020/05/12; 2021/07/15 |
| Saudi Arabia | B | 3 | 412 | 2020/06/20<br>2021/08/06 | 2021/07/02 |
| South Africa | B/C | 4 | 178 | 2021/01/11<br>2021/07/08 | 2020/07/19; 2021/08/22 |
| South Korea | C/D | 7 | 46 | 2021/08/15<br>2021/09/30 | 2020/03/04; 2020/08/27; 2020/12/25;<br>2021/02/20; 2021/04/23 |
| Spain | B/C | 5 | 174 | 2021/01/26<br>2021/07/19 | 2020/03/31; 2020/11/04; 2021/04/27 |
| Sweden | D/E | 4 | 91 | 2021/01/11<br>2021/04/12 | 2020/04/29; 2020/06/18 |
| Turkey | B/C/D | 5 | 139 | 2020/12/02<br>2021/04/20 | 2020/04/16; 2021/08/15; 2021/10/15 |
| UK | C | 6 | 193 | 2021/01/09<br>2021/07/21 | 2020/04/22; 2020/11/16; 2021/09/08;<br>2021/10/23 |
| USA | A/B/C/D/E | 5 | 245 | 2021/01/11<br>2021/09/13 | 2020/04/10; 2020/07/22; 2021/04/14 |

Precisely, the number of peaks, the distance in days between the two highest ones and their corresponding dates are given for each country. It is interesting to notice that if we average, all over the 30 countries, the values of the temporal distances between the two highest peaks, we obtain a mean of 190 days (SD 100). In other words, we have obtained a confirmation for all our 30 countries of the recurrence of peaks, with an average period of almost 6 months and a standard deviation of nearly 3 months. Moreover, the 80% of the examined countries has that (maximal) temporal distance which falls below the value of one year. Even if we restrict this analysis to only the 13 European countries of our dataset: Austria, Belgium, Croatia, Denmark, France, Germany, Hungary, Italy, Norway, Portugal, Spain, Sweden, and the UK, we achieve an average of almost 5 peaks in a two-years period, with a mean distance between the two highest ones equal to 171 days (SD 85), once again confirming the hypothesis that strong COVID-19 waves may repeat with cycles whose duration break the seasonality pattern of one year.

## Discussion

Seasonality typically refers to a single, recurring pattern with a fixed frequency. The results we achieved highlight how, with COVID-19, strong evidence of a seasonal pattern that repeats over a one-year fixed period cannot be found. Instead, several repeating outbreaks, not necessarily occurring with a fixed frequency, can be observed. In particular, the Fourier spectral analysis of the time series of the SARS-CoV-2 cases of all the 30 countries we have studied has revealed the recurrence of at least one COVID-19 wave (often two or more), repeating over a variable period, in the range 3 - 9 months, with peaks of magnitude comparable to that of the highest peak ever experienced since the beginning of the pandemic until December 2021. Indeed, the situation is more complicated than previous studies have revealed. In fact, while some of them consider COVID-19 as a seasonal low-temperature infection, the precise role of temperature, humidity, and exposition to UV radiation remains poorly understood. From this perspective, our study has tried to follow an alternative path with spectral analysis of the series of the daily SARS-CoV-2 cases, while looking for peaks in the frequency spectrum to understand the presence of repeating cycles and quantify their lengths.

We recognize that, with our analysis, we have avoided quantifying precisely the role attributed to various climatic factors or control measures, like temperatures or vaccination. This can be seen as a limitation of our study as the magnitude of the effects of those factors should be investigated thoroughly. Yet, there are precise motivations behind our choice. On one side, we have decided to avoid taking part in the scientific discussion about the dominant role of climate vs. control measures, including vaccination, as the best solutions that can drive substantial changes to the pandemic trajectory. On the other side, we have tried to observe a natural phenomenon, just resorting to a mathematical technique able to detect the presence of evidence towards periodicity/non-periodicity in the spread of COVID-19, with neutrality and regardless of the underlying factors.

Another technical limitation of our approach was the decision not to put a special focus on those countries where the number of cases has had high variability, the reason being most likely that the number of tests done each day can have varied as much. We could have normalized those cases with the number of tests, before subjecting them to the discrete Fourier transform, but this datum is often unreliable and may lead, in turn to unrealistic normalized values, so we decided to avoid this. An additional technical limitation of this study is that the Fourier transform may return results, especially at the lowest frequencies, with a variable degree of uncertainty. Hence, to confirm our results, we have developed a parallel analysis directly performed on the number of the new daily SARS-COV-2 cases of interest. Alternatively, we could have performed a spectral analysis of our epidemiological time-series with wavelets. These techniques appear somewhat attractive because they are more appropriate to treat non-stationary signals, but still have to deal with the natural limitation represented by the fact that the pandemic has been in progress so far, and we only have a two-year dataset. Finally, a limitation of our study refers to the exclusion of the intense SARS-CoV-2 outbreak, which started in December 2021 in many of the 30 considered countries. The motivation for this exclusion is that the progression of this wave is unfortunately still ongoing.

## Conclusion

We have applied a mathematical technique from signal processing to investigate in 30 different countries the hypothesis whether COVID-19 outbreaks either repeat with a fixed periodicity or occur following unpredictable patterns. The Fourier spectral analysis of the time series of the SARS-CoV-2 cases we have examined has suggested that strong COVID-19 waves may repeat with cycles of different lengths, without a precisely predictable periodicity (one year, or similar). With the scientific community that appears divided into two factions, which alternatively maintain the importance of the role of meteorological factors vs. control measures (including vaccination), we argue we have provided an improved understanding of how the virus may spread, regardless of the presence of several factors that can further confound the scenario.

## Data Availability

Data are available in a public, open access repository (https://github.com/owid/covid-19-data). All data and code are also available upon request to the corresponding author

## Author Contribution

FC, DT, LC, and MR equally contributed to conceive, design, write, manage, and revise the manuscript.

## Patient consent for publication

Not required.

## Provenance and peer review

Not commissioned, externally peer reviewed.

## Data availability statement

Data are available in a public, open access repository (https://github.com/owid/covid-19-data). All data and code are available upon request to the corresponding author

## Research Ethics Approval: Human Participants

Not applicable: neither humans nor animals nor personal data are being involved in this study.

